# MID-LIFE LEISURE-TIME PHYSICAL ACTIVITY IS ASSOCIATED WITH LOWER HIP FRACTURE RISK IN UK BIOBANK

**DOI:** 10.64898/2026.10.08.26365023

**Authors:** Catherine Rolls, Benjamin G. Faber, Emma M Clark, Helen Dawes, Marcus R Munafò, Jonathan H Tobias

## Abstract

Leisure time physical activity (LTPA) in mid-life may have beneficial effects on risk factors for hip fracture in later life. Conversely, physical activity may increase fracture risk through risk of injury, and the optimal amount of physical activity for reducing hip fracture risk remains unclear. We explored the associations between volume and intensity of LTPA in mid-life, and subsequent risk of hip fracture ascertained from linked health data. This prospective observational study used middle aged adults aged 40-65 years from UK Biobank. LTPA volume (LTPAv) was derived from self-reported walking, moderate and vigorous activity at baseline (2006-2010). Cox proportional hazard and Fine-Gray sub-distribution models were used to estimate the association between activity and hip fracture, which was sex-stratified. From 324,284 participants with complete data 53% were female and mean age at baseline was 54.5 (SD 7.3). Across 16 years of follow-up there were 2,674 hip fractures. In terms of LTPA volume, the lowest risk of hip fracture occurred in quintile 3 for women (HR 0.78, 95% CI 0.67–0.91), and men (HR 0.74, 95% CI 0.60–0.90), relating to approximately one hour/day of leisure time physical activity. When activity intensities were examined separately, the lowest estimated hazards were observed for vigorous activity, at quintile 4 in women (HR 0.65, 95% CI 0.55–0.76) and quintile 3 in men (HR 0.64, 95% CI 0.52–0.79), corresponding to approximately 60–90 minutes of vigorous activity per week. In conclusion, moderately high volumes of mid-life physical activity, particularly vigorous activity, were related to lower risk of subsequent hip fracture, but these relationships appear to attenuate at higher volumes.

**Lay Summary:** It is not clear whether being active in mid-life, aged 40-65 years, reduces your risk of having a hip fracture in older age. We used data from 324,284 people from the UK Biobank, a long running health study, to understand this better. We found that people who were more active had a lower risk of hip fracture. Risk was lowest among people doing about an hour a day of leisure-time physical activity overall, but less benefit was observed with higher amounts. When we looked at different activity intensities, those doing about 60-90 minutes a week of more vigorous activities like running had the lowest risk. This suggests that both the amount and intensity of activity matters.

## INTRODUCTION

Promoting healthy ageing has become a national policy priority in the UK, with a focus on enabling people to live *“an extra 5 years of healthy, independent life by 2035”*.^1^ Hip fractures are one of the most important age-related conditions that contribute to loss of independence, functional decline and loss of life. In the UK alone over 70,000 adults sustain a hip fracture annually, and cumulative costs following a hip fracture were over £869 million in the last 10 years.^2^ Mortality within a month of injury is high and for those who survive only half regain previous levels of mobility resulting in a loss of independence and an increase in institutionalisation.^3,4^ While effective treatments exist for osteoporosis, a common cause of hip fracture, these are typically initiated after individuals have been identified as being at high risk or following a fracture. Consequently, strategies that reduce fracture risk before a first fracture occurs remain a public health priority.

Physical activity is a potentially cost-effective fracture prevention strategy, with biologically plausible mechanisms through improved bone and muscle strength, balance and reduced falls. Because hip fracture is relatively rare, intervention trials have commonly used Bone Mineral Density (BMD) as a surrogate outcome with current evidence suggesting that high intensity resistance and impact training can lead to a 2-3% increase in BMD at the femoral neck.^5^ However, the strongest effects are seen in supervised interventions and post-menopausal women with low BMD.^5,6^ In addition sustaining prescribed exercise over the long term is difficult, with higher intensity interventions and studies of longer duration having significantly higher rates of non-compliance.^7^ Evidence from qualitative research suggests that individuals are more likely to maintain physical activity if it is self-selected and done for pleasure, with less certainty in relation to exercise for health benefits alone.^8^ This raises the question of whether habitual leisure time physical activity could offer similar benefits for bone health to supervised interventions, and highlights the need for studies with long term follow-up.

Despite strong mechanistic evidence for bone adaptation, results from observational studies on the effects of leisure time physical activity (LTPA) on hip fracture risk are mixed.^9^ There is moderate evidence that women who are highly active have a reduced risk of future hip fracture, but the evidence for a protective effect of moderate activity, or in men is less convincing.^9^ A recent review identified considerable variation in the measurement and reporting of physical activity, with studies differing in the questionnaires used, the quantification of exercise, and the definition of the exposure.^9^

Mid-life may represent a particularly important stage for intervention because individuals are generally more capable of undertaking higher-intensity and higher impact exercise before the decline in bone mass, muscle function and mobility that accompanies older age. While physical activity in mid-life has well established long-term benefits for cardiovascular disease and cancer,^10,11^ the evidence for hip fracture is more nuanced. Physical activity has the potential for increased exposure to falls during exercise and higher-energy trauma. Although the available evidence suggests that physical activity during mid-life may reduce subsequent hip fracture risk, it remains unclear whether this benefit varies according to the volume or intensity of activity undertaken. This is particularly relevant given recent cross-sectional evidence from our group that relatively high volumes of LTPA and vigorous activity were associated with higher fracture risk in mid-life.^12^

Existing evidence from prospective cohort studies with fracture end points on the long term benefits and risks of activity participation remains uncertain. There is a paucity of data on the optimal range for volume and intensity in mid-life. We examined the prospective associations between mid-life physical activity volume and intensity and subsequent risk of hip fracture in UK Biobank (UKB), which provides up to 16 years of follow-up.

## MATERIALS AND METHODS

### Study design and population

This study is a prospective observational analysis using self-reported activity data and hip fracture derived from linked hospital records in UKB, a large, long-running cohort study that recruited around 500,000 adults across the United Kingdom between 2006 and 2010. It comprises comprehensive self-reported health and lifestyle information alongside objectively measured data, including height, weight and bone mineral density (BMD). Most participants consented to linkage with electronic health records, enabling ascertainment of hospital admissions through the Hospital Episode Statistic (HES) database.^13^ While the age of the cohort at recruitment was 40-69, for the purpose of this study we restricted our analysis to middle aged adults (40-65 years at recruitment).

### Leisure time physical activity

Our exposure was LTPA, derived from self-reported activity at the baseline assessment visit in 2006-2010. UKB use a modified version of the validated International Physical Activity Questionnaire (IPAQ)^14^ to ask about LTPA undertaken in the previous week. Activity volume (minutes/week) was calculated from frequency (days/week) and duration (minutes/day) of walking for pleasure, moderate intensity, or vigorous intensity activity. Metabolic equivalent task (MET)-minutes/week were used to capture both the volume and intensity of activity. These were calculated using standard intensity weights multiplied by weekly volume.^15^ We derived separate MET-minutes/week variables for walking, moderate and vigorous activity. LTPA volume was calculated as the sum of all components, expressed from here on as LTPA_V_.

### Fracture ascertainment

The primary outcome was incident hip fracture. Hip fractures were defined using ICD-10 codes S72.0-72.2. The complete list of ICD-10 codes with descriptions are provided in Supplementary Table S1. Hospital admissions and mortality data were obtained through linked national health records.

### Covariates

Potential confounders of the relationship between mid-life physical activity and fracture risk were identified a priori using a directed acyclic graph (Supplementary Figure S1) from which we developed a minimally sufficient adjustment set. Sex and ethnicity were taken from the baseline assessment visit. Sex was defined as male or female, while ethnicity was self-reported and categorised as White, Asian, British Asian, Black, Black British, or mixed ethnicity. Educational attainment was grouped as school education or less, college or vocational training, and university degree or above. We used the Townsend Deprivation Index as the measure of socioeconomic deprivation, where a higher score indicates greater deprivation.^16^ Standing weight and height were measured at the baseline assessment centre visit according to published protocols.^17^

### Statistical analysis

This study is reported in accordance with the Strengthening the Reporting of Observational Studies in Epidemiology guidelines.^18^ The analysis plan for the study was pre-registered,^19^ and minor deviations from the study protocol are documented in Supplementary Methods S1. Code is available at Github (https://github.com/RollsCf/UKB_PA_Fracture.git.)

Continuous variables are presented as mean (standard deviation, SD) or median (Interquartile range, IQR). Categorical variables are described as frequencies and percentages. We restricted analysis to participants with complete data for the exposure and covariates, and who consented to hospital data linkage. Baseline characteristics of included and excluded participants were compared using standardised mean differences.

Cox proportional hazard regression was used to estimate hazard ratios (HRs) and 95% confidence intervals (CIs) for incident fracture, the proportional hazards assumption was checked with Schoenfeld residuals. Time-to-event analyses used attained age as the timescale as such we did not also adjust for age. Participants entered the risk set at their age at study entry and were followed until the age at first hip fracture or censored at the earliest of death, loss to follow-up - defined as withdrawal from UKB, or the end of available linked hospital records. The end of follow-up differed by country according to data availability (England: 31 March 2023; Scotland: 31 August 2022; Wales: 31 May 2022).

Three models were fitted with progressive levels of adjustment: (1) unadjusted; (2) minimally adjusted for sex, ethnicity, weight and height; and (3) fully adjusted, additionally including socioeconomic deprivation and education. Physical activity variables were right-skewed and were therefore log-transformed prior to modelling (log [MET+1]). Non-linearity in the association between physical activity and fracture risk was assessed using restricted cubic splines with four knots, with hazard ratios estimated relative to the median level of physical activity. The proportional hazards assumption was assessed for all Cox models. Effect modification by sex was evaluated by including interaction terms, with likelihood ratio tests (LRT) comparing models with and without interactions terms. To aid interpretation, physical activity was also modelled as quintiles of the original (untransformed) exposure, with the lowest quintile (Q1) as the reference group. To examine associations with different activity intensities, walking, moderate and vigorous activity volumes (MET-minutes/week) were modelled simultaneously, such that associations for each intensity were adjusted for the other activity intensities.

To assess the robustness of our findings, we conducted further sensitivity and exploratory analyses. First, Fine–Gray competing-risks models were fitted to account for death as a competing event. Second, to examine whether the observed associations were independent of baseline skeletal and physical-function characteristics, we conducted additional exploratory analyses. The models were separately extended to include baseline heel BMD, derived from heel ultrasound, and established measures of physical function, comprising grip strength, self-reported walking speed and history of falls.^20,21^ All analysis were conducted with R (version 4.5.2).

## RESULTS

### Population characteristics

There were 324,284 people with complete data, with a mean of 13.7 year of follow-up (SD 1.9; maximum 16.2 years. Fifty three percent were female, the average age was 54.5 (SD 7.3), 95% of the cohort were of White ethnicity (Table 1). Missingness was low for most variables (<5%), although higher for measures of activity with the composite measure of LTPA having 22% missingness. Those excluded for having missing data at baseline had comparable baseline characteristics to those included (Supplementary Table S2-3). The flow of participants through the study is documented in Fig 1.

**Figure 1:**
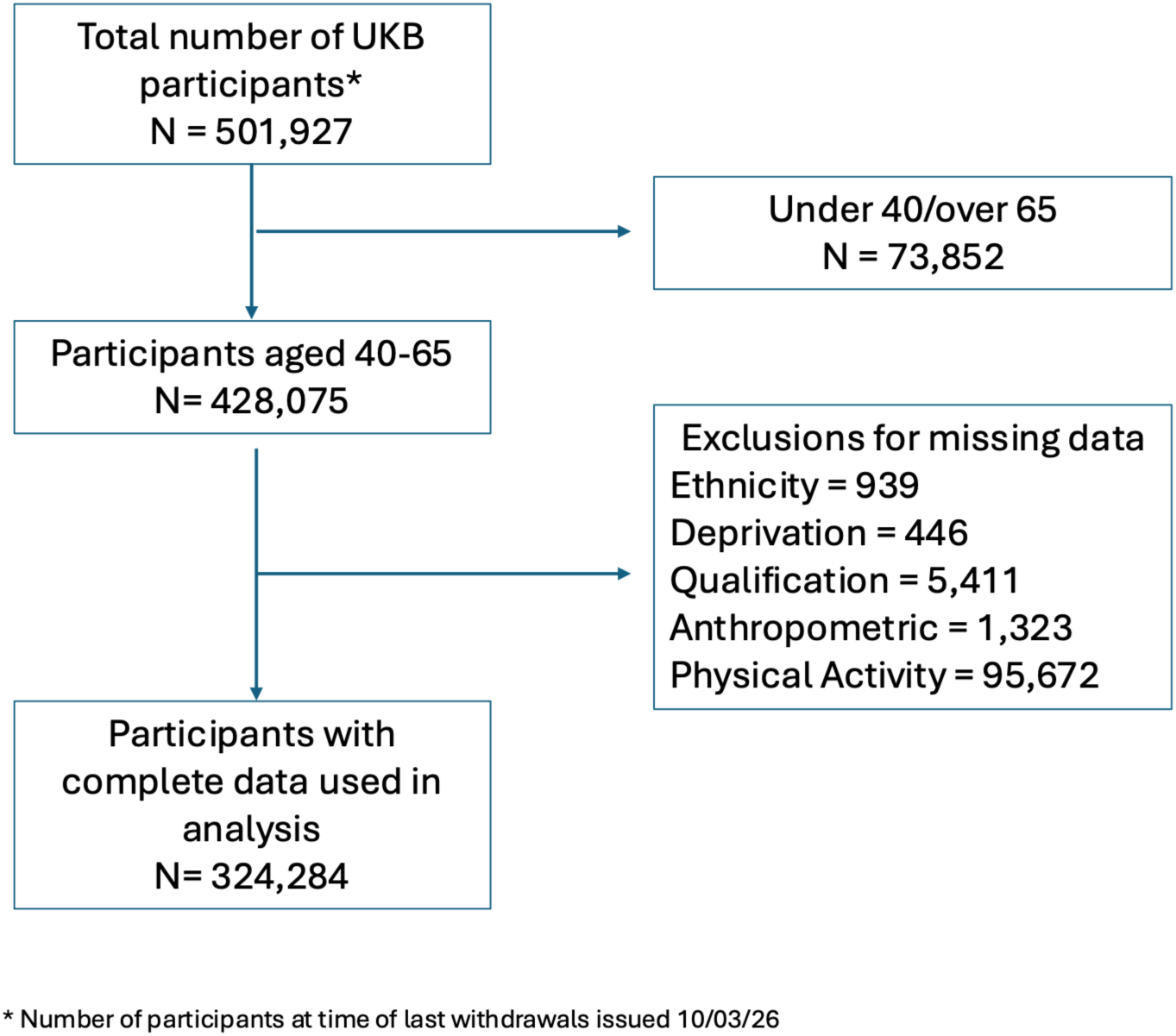
**Flow diagram of participant selection for the study**. Flow diagram showing the selection of UK Biobank participants included in the analyses of leisure time physical activity (LTPA) and hip fracture. Participants were excluded sequentially according to the criteria shown. The final analytical cohort comprised participants aged 40–65 years with complete data for the exposure, outcome and covariates included in the analyses.

**Table 1:** Demographics Table.

| Variable | Sex |  |  |
| --- | --- | --- | --- |
|  | Overall N = 324,284 | Female N = 171,699 | Male N = 152,585 |
| Age (years), mean (SD) | 54.5 (7.3) | 54.3 (7.2) | 54.7 (7.3) |
| Ethnicity, n (%) |  |  |  |
| Asian | 5,642 (1.7%) | 2,578 (1.5%) | 3,064 (2.0%) |
| Black or other | 8,804 (2.7%) | 5,040 (2.9%) | 3,764 (2.5%) |
| Mixed | 2,058 (0.6%) | 1,282 (0.7%) | 776 (0.5%) |
| White | 307,780 (95%) | 162,799 (95%) | 144,981 (95%) |
| Townsend deprivation index, mean (SD) | -1.4 (3.0) | -1.4 (3.0) | -1.4 (3.1) |
| Education level, n (%) |  |  |  |
| School education or less | 272,378 (84%) | 144,731 (84%) | 127,647 (84%) |
| College or vocational training | 30,909 (9.5%) | 15,506 (9.0%) | 15,403 (10%) |
| University degree or above | 20,997 (6.5%) | 11,462 (6.7%) | 9,535 (6.2%) |
| Weight (kg), mean (SD) | 78.3 (16.0) | 71.2 (14.0) | 86.2 (14.3) |
| Height (cm), mean (SD) | 169.2 (9.3) | 163.0 (6.3) | 176.2 (6.8) |
| Follow-up (years), mean (SD) | 13.7 (1.9) | 13.8 (1.8) | 13.6 (2.1) |
| Hip fracture, n (%) | 2,674 (0.8%) | 1,689 (1.0%) | 985 (0.6%) |
| Moderate activity (MET-min/week), median (IQR) | 480 (120, 1,200) | 480 (120, 1,200) | 420 (120, 1,200) |
| Vigorous activity (MET-min/week), median (IQR) | 240 (0, 960) | 192 (0, 720) | 320 (0, 1,000) |
| Walking (MET-min/week), median (IQR) | 693 (297, 1,386) | 693 (330, 1,386) | 693 (297, 1,386) |
| Leisure time physical activity (MET-min/week), median (IQR) | 1,771 (813, 3,546) | 1,733 (810, 3,375) | 1,818 (813, 3,732) |

Continuous demographic and anthropometric variables and follow-up are presented as mean (standard deviation (SD)). Physical activity variables are presented as median (inter quartile range (IQR)). Categorical variables are presented as n/N (%). MET = metabolic equivalent Task unit

### Distributions and characteristics of physical activity and fracture in mid-life

Physical activity distributions were right skewed across all domains and were subsequently log transformed (log +1) for modelling (Supplementary Figure S2). Men reported higher LTPA_V_ than women with a median of 1,818 (IQR 813-3732) compared to 1,733 (IQR 810-3378) MET-min/week. Walking for pleasure made up the highest proportion of activity for men and women. Women reported a higher proportion of moderate activity (median of 480 MET-min/week (IQR 120-1200) compared to 420 (IQR 120-1200), with men more likely to report participation in vigorous activity (median 320 compared to 192 MET-min/week (IQR 0-1000). Across up to 16 years of follow-up there were 2,674 hip fractures, with 63% sustained by women (Table 1).

### LTPA_V_ and fracture risk

Cox proportional hazards model revealed an inverse relationship between LTPA_V_ and risk of hip fracture. This association was strengthened by minimal adjustment for sex, ethnicity, weight and height, with little further change after additional adjustment for socioeconomic deprivation and education. Likelihood ratio testing demonstrated evidence of non-linearity (p for non-linearity <0.001); therefore, a restricted cubic spline model was used to characterise the dose response relationship. Tests of Cox proportional hazards model for linearity and interaction are presented in Supplementary Table S4. There was no evidence of effect modification by sex, however, the proportional hazards assumption was violated, with sex contributing substantially to the violation (Supplementary Table S5). Consequently, subsequent analyses of LTPA_V_ were stratified by sex. Remodelling demonstrated all proportional hazards assumptions were met.

In the sex-stratified restricted cubic spline model, greater LTPA_V_ was associated with a lower risk of subsequent hip fracture, with a similar pattern in males and females (Figure 2). Hip fracture hazard declined with increasing LTPA_V_, reaching its lowest level at approximately 1,800 MET-min/week (back transformed from the logged units). Beyond this point, the estimated hazard increased modestly, indicating attenuation of the inverse association at higher activity volumes; however, confidence intervals widened at the highest activity levels.

**Figure 2.**
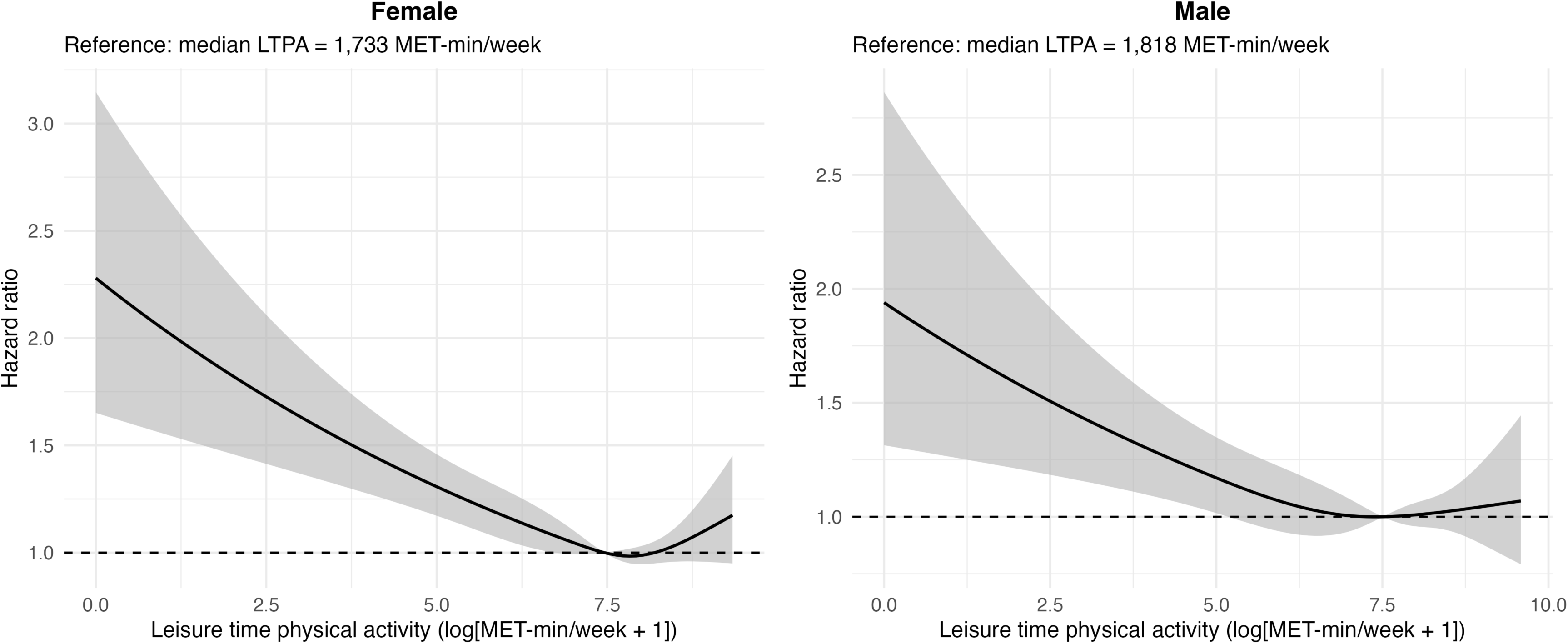
**Restricted cubic spline associations between total leisure-time physical activity and the risk of hip fracture, stratified by sex**. Total physical activity was modelled as log(MET-min/week + 1). Solid lines represent adjusted hazard ratios (HRs), and shaded areas represent 95% confidence intervals. Hazard ratios are presented relative to the median level of total physical activity (HR = 1.0; dashed horizontal line). Models were adjusted for age, ethnicity, socioeconomic deprivation, smoking status, alcohol consumption, body size, comorbidity, menopausal status (women), and prior physical activity.

To facilitate interpretation of the non-linear dose-response relationship, LTPA_V_ was categorised into sex-specific quintiles (Q). Table 2 summarises the activity volumes represented within each quintile. The median activity volume in quintile 3 was 1,770 MET-min/week for both men and women, equivalent to around 420 minutes of activity per week (approximately one hour per day), closely corresponding to the plateau observed in the restricted cubic spline models.

**Table 2.** Activity composition and selected musculoskeletal characteristics across physical activity quintiles by sex.

| Characteristic | Statistic | Q1 | Q2 | Q3 | Q4 | Q5 |
| --- | --- | --- | --- | --- | --- | --- |
| Female |  |  |  |  |  |  |
| LTPA, MET-hours/week | Median (IQR) | 5.8 (3.3–8.2) | 16.3 (13.5–19.3) | 29.5 (25.6–33.7) | 51.1 (44.0–59.1) | 99.3 (82.2–129.3) |
| LTPA, hours/week | Median (IQR) | 1.7 (0.8–2.3) | 4.0 (3.5–5.0) | 7.1 (6.2–8.3) | 12.0 (10.3–14.5) | 24.5 (20.2–31.5) |
| Walking, hours/week | Median (IQR) | 1.0 (0.5–1.7) | 2.5 (1.5–3.5) | 3.5 (2.1–6.0) | 5.8 (3.5–8.8) | 14.0 (7.0–21.0) |
| Moderate activity, hours/week | Median (IQR) | 0.2 (0.0–0.5) | 1.0 (0.5–1.8) | 2.0 (1.0–3.0) | 4.0 (2.0–6.0) | 10.0 (6.0–15.0) |
| Vigorous activity, hours/week | Median (IQR) | 0.0 (0.0–0.0) | 0.2 (0.0–0.7) | 0.7 (0.0–1.5) | 1.5 (0.2–2.7) | 2.5 (0.7–5.0) |
| Brisk walking pace | n (% across quintiles) | 9,732 (13.1%) | 14,301 (19.3%) | 16,076 (21.7%) | 17,346 (23.4%) | 16,655 (22.5%) |
| More than one fall | n (% across quintiles) | 2,680 (24.0%) | 2,074 (18.5%) | 2,007 (17.9%) | 2,128 (19.0%) | 2,295 (20.5%) |
| Grip strength, kg | Median (IQR) | 23.5 (19.5–28.0) | 24.0 (20.0–28.0) | 24.0 (20.0–28.5) | 24.5 (20.0–28.5) | 24.5 (20.0–29.0) |
| Heel BMD | Median (IQR) | 0.51 (0.44–0.59) | 0.51 (0.44–0.59) | 0.52 (0.45–0.60) | 0.52 (0.44–0.60) | 0.52 (0.44–0.60) |
| Male |  |  |  |  |  |  |
| LTPA, MET-hours/week | Median (IQR) | 5.5 (2.5–8.2) | 16.4 (13.6–19.3) | 29.5 (25.7–33.8) | 51.0 (44.0–59.1) | 107.1 (85.1–147.0) |
| LTPA, hours/week | Median (IQR) | 1.5 (0.7–2.2) | 4.0 (3.4–4.8) | 7.0 (6.0–8.1) | 11.8 (9.9–14.0) | 25.8 (20.2–35.0) |
| Walking, hours/week | Median (IQR) | 1.0 (0.3–1.7) | 2.5 (1.3–3.5) | 3.5 (2.0–6.0) | 5.2 (2.9–7.0) | 14.0 (7.0–21.0) |
| Moderate activity, hours/week | Median (IQR) | 0.2 (0.0–0.5) | 1.0 (0.3–1.5) | 2.0 (1.0–3.0) | 3.5 (2.0–5.2) | 10.5 (6.0–15.0) |
| Vigorous activity, hours/week | Median (IQR) | 0.0 (0.0–0.0) | 0.2 (0.0–0.8) | 1.0 (0.0–2.0) | 2.0 (0.5–3.0) | 3.5 (1.2–6.3) |
| Brisk walking pace | n (% across quintiles) | 10,757 (16.2%) | 12,861 (19.3%) | 13,781 (20.7%) | 14,248 (21.4%) | 14,840 (22.3%) |
| More than one fall | n (% across quintiles) | 1,859 (23.8%) | 1,184 (15.2%) | 1,199 (15.4%) | 1,383 (17.7%) | 2,185 (28.0%) |
| Grip strength, kg | Median (IQR) | 39.5 (33.5–45.5) | 40.0 (34.5–46.0) | 40.0 (35.0–46.0) | 41.0 (35.0–46.5) | 41.0 (35.5–47.0) |
| Heel BMD | Median (IQR) | 0.55 (0.47–0.63) | 0.56 (0.48–0.64) | 0.56 (0.49–0.65) | 0.57 (0.49–0.66) | 0.57 (0.49–0.66) |

Values are median (interquartile range (IQR)) for continuous variables and n (%) for categorical variables. MET-hours/week were calculated as MET-min/week divided by 60. Q = Quintile, LTPA = Leisure Time Physical Activity, kg = kilograms. MET = Metabolic Task Equivalent, BMD = Bone Mineral Density, Kg = kilogram

Associations between LTPA_V_ quintiles and hip fracture were examined using Cox proportional hazards models (Figure 3). In fully adjusted models, women demonstrated the lowest risk of hip fracture in Q3 (HR 0.78, 95% CI 0.67–0.91), with smaller effect estimates in Q4 and 5, suggesting little additional reduction in risk at higher activity volumes. Among men, the greatest reduction in hip fracture risk also occurred in Q3 (HR 0.74, 95% CI 0.60–0.90), whereas associations for Q4 and 5 overlapped with the null. Full model estimates are presented in Supplementary Table S6.

**Figure 3.**
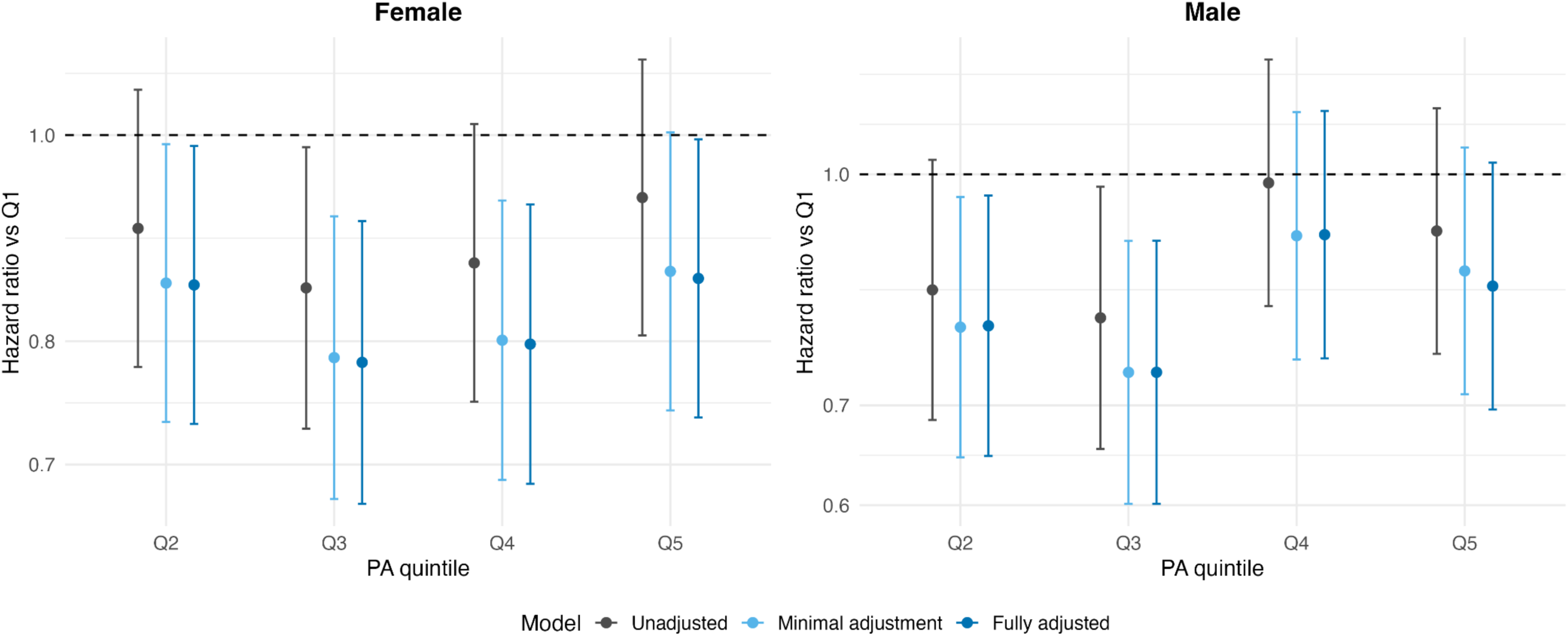
**Hazard ratios (HRs) and 95% confidence intervals for hip fracture according to quintiles of total leisure-time physical activity, stratified by sex**. Estimates are shown from unadjusted, minimally adjusted, and fully adjusted Cox proportional hazards models. Quintile 1 (lowest physical activity) was used as the reference category (HR = 1.0). Error bars represent 95% confidence intervals.

Fine-Gray models accounting for death as a competing event demonstrated dose-response relationships that were highly consistent with the primary Cox models, with only modest attenuation of the effect estimate. (Supplementary Figure S3).

### Activity intensity and fracture risk

The composition of activity was broadly comparable between men and women across the quintiles, with participants in quintile one undertaking on average 1 hour a week of walking activity and little to no moderate or vigorous activity. Composition remained similar in quintile three with an average of 3.5 hours a week of walking, 2 of moderate and 0.7-1 hour of vigorous behaviour. In quintile five, while walking and moderate activity behaviours remained similar between the sexes with an average of 14 hours/week to 10 hours per week respectively, men undertook more vigorous activity (3.5 hours/week (SD 1.2-6.3), compared to 2.5 hours/week (SD 0.7-5.0) in women (Table 2).

We examined associations between quintiles of walking, moderate (not including walking) and vigorous activity and hip fracture risk, in mutually adjusted models. Associations differed by intensity and sex (Figure 4). In women walking was not associated with a change in fracture risk, while moderate activity was associated with reduced fracture risk across Q2-4. The lowest estimated hazard was observed for Q4 of vigorous activity (HR 0.65, 95% CI 0.55-0.76). In men, walking and moderate activity did not appear to be associated with consistent reduction in fracture risk. The lowest estimated hazard was observed for vigorous activity in Q3 (HR 0.64, 95% CI 0.52-0.79), corresponding to approximately 60-90 minutes of vigorous activity per week. Full model estimates are presented in Supplementary Table S7.

**Figure 4.**
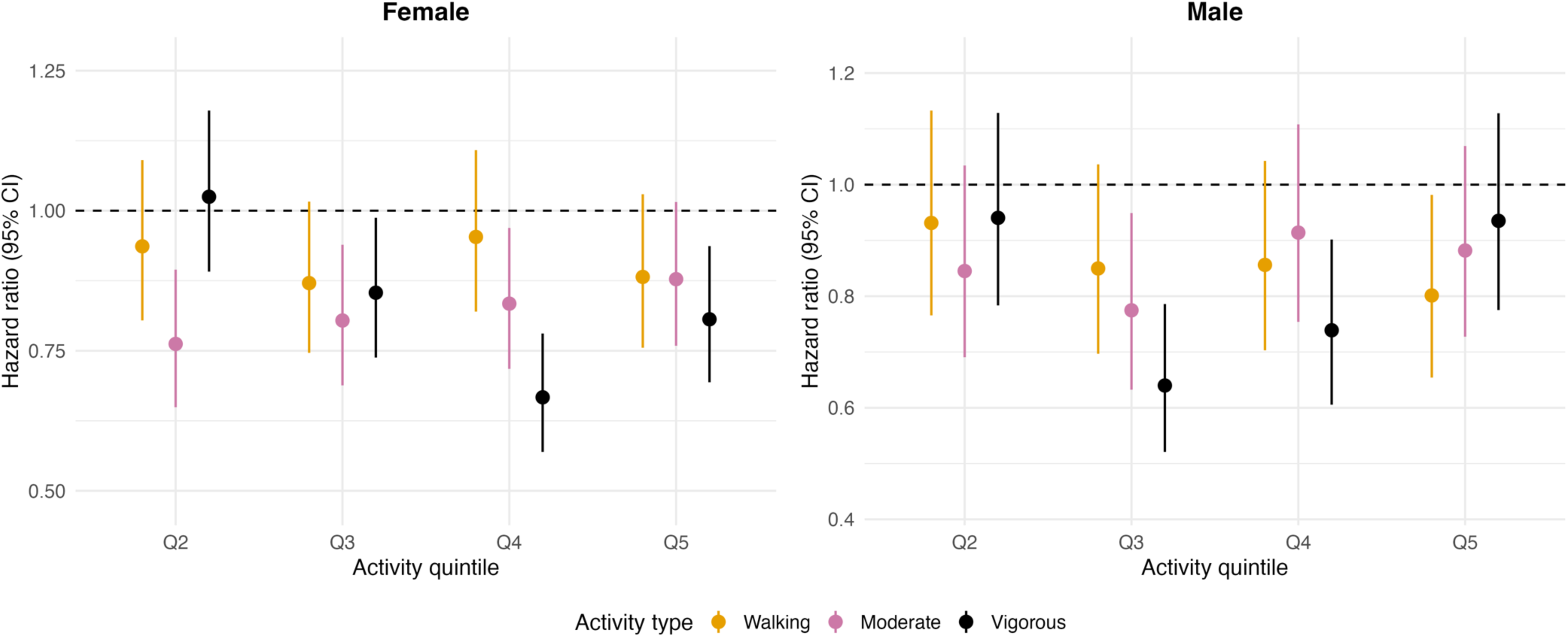
Hazard ratios (HRs) and 95% confidence intervals for hip fracture according to quintiles of walking, moderate-intensity, and vigorous-intensity physical activity, stratified by sex. Estimates are from fully adjusted Cox proportional hazards models with walking, moderate-intensity, and vigorous-intensity physical activity included simultaneously to estimate the independent association of each activity type with fracture risk. Quintile 1 (lowest activity level) was used as the reference category

### Exploration of further models relating PA and fracture risk

We explored the role of other possible confounders/mediators of the relationship between LTPA and hip fracture risk. Indicators of physical function and baseline heel BMD by activity quintile and sex are reported in Table 2. Grip strength was almost double in men compared to women but did not differ notable by activity level. Differences in baseline heel BMD across activity quintiles were small. Falls history appeared to have a U-shaped pattern with highest number of participants having experienced more than one fall being in Q1 and 5. Self-reported walking pace increased with increasing activity levels in men and women, before plateauing at Q4 in women.

When our adjustment model for LTPA_V_ was extended to include grip strength, falls history and heel BMD separately, these had little material effect on the hazard ratios (Figure 5), with similar findings across all other activity exposures. By contrast, additional adjustment for self-reported walking pace attenuated the effect estimates towards the null across all exposures, although the strongest inverse associations between vigorous activity and hip fracture risk persisted (Q3 in men, Q4 in women) (Supplementary Figure S4). Results for full analyses are presented in Supplementary Table S8.

**Figure 5:**
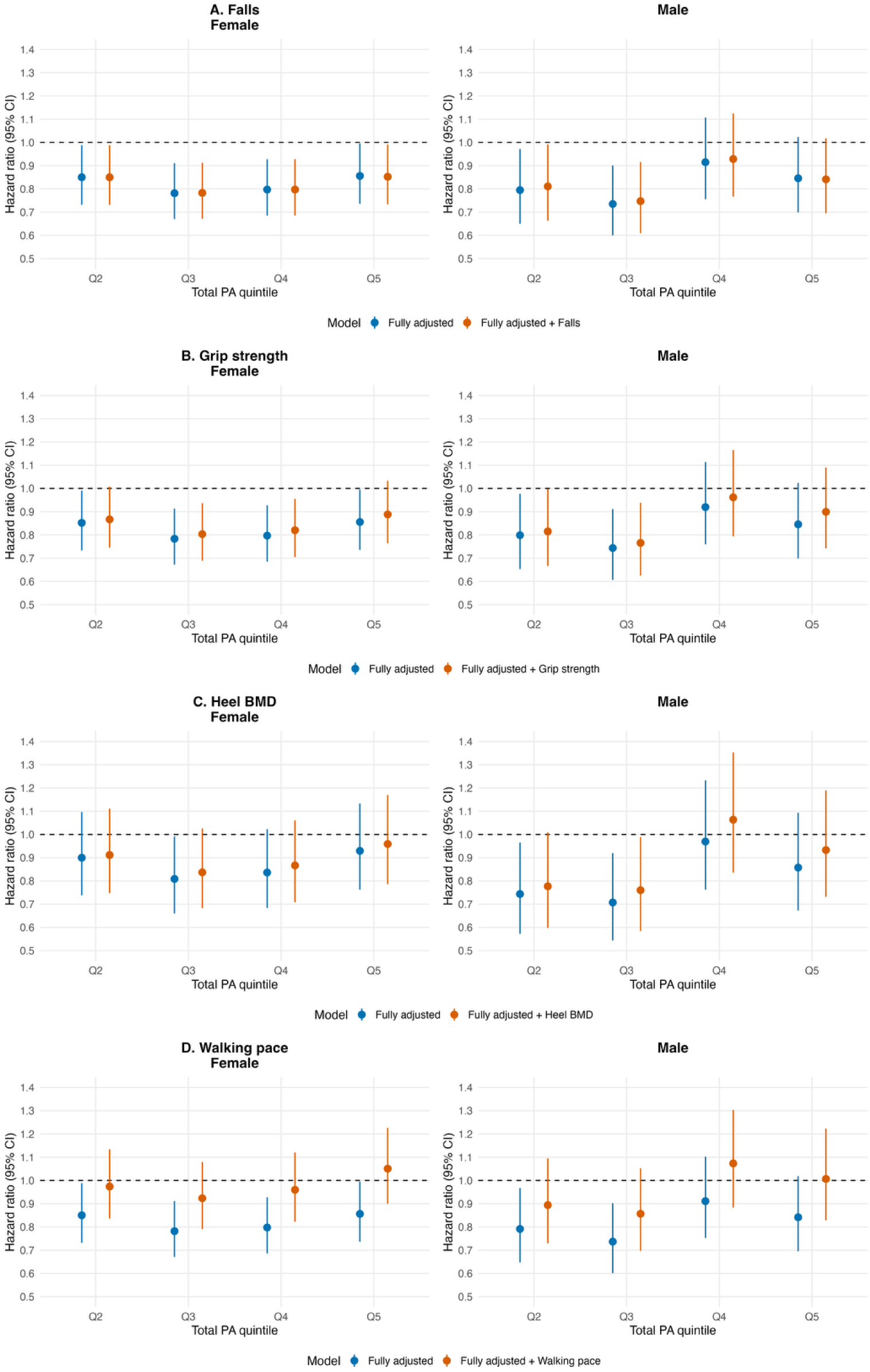
Cox proportional hazard models for LTPAv and hip fracture extended to adjust for indicators of physical function. Hazard Ratios (95% confidence intervals) are shown for activity quintiles Q2-Q5 relative to the lowest activity quintiles (Q1; reference category). Estimates are presented as fully adjusted for ethnicity, height, weight, Townsend deprivation index and education level and fully adjusted plus either Grip strength, baseline heel BMD, or self-reported walking pace

## DISCUSSION

We examined the prospective associations between LTPA_V_ and the amount of walking, moderate and vigorous activity undertaken in mid-life and risk of future hip fracture. Higher volumes of LTPA were associated with lower hip fracture risk, with the lowest risk observed at approximately one hour of total LTPA per day. For vigorous activity, approximately 60-90 minutes per week was associated with the lowest risk, independent of walking and moderate activity. The inverse associations attenuated at higher activity volumes, suggesting that the relationship between mid-life physical activity and later hip fracture risk is non-linear and that both the volume and intensity of activity may be important.

A key finding from this study is that an increase in LTPA_V_ was associated with lower hip fracture risk, with the lowest estimated hazard at approximately 1800 MET minutes/week (approximately 420 mins per week (1 hour per day) of LTPA) and slight attenuation at higher volumes. The attenuation of effect at the highest volume of activity was mirrored in the quintile analyses with the greatest reduction in hazard of hip fracture occurring in quintile 3. Secondly, our study suggests not only volume of LTPA matters but also how it is accumulated. Previous observational research into activity and hip fracture risk has not been able to separate the role of total activity volume from intensity. We found that an increase in vigorous intensity activity, equating to around 60-90 minutes a week, had the strongest inverse association with future hip fracture risk independent of walking and moderate intensity activity. Consistent with the findings for overall LTPA_V_, the inverse association attenuated slightly at highest volumes of vigorous activity.

A possible explanation for the slight attenuation in hip fracture reduction at highest activity levels is suggested by our previous cross-sectional study, which found that at the highest volume and intensity of LTPA odds of fracture were increased, potentially due to the increased exposure to falls and trauma that arise from regular participation in sporting activities that contribute to vigorous activity.^12^ In the present study, since activity was only measured at baseline, subsequent trajectory of activity up to the time of hip fracture is unknown. Nevertheless, to the extent that those with highest levels of activity at baseline are may be more likely to partake in activities with greater risk of injury in later life, this might explain the partial attenuation of the association between greater activity and reduced risk of hip fracture that we observed. Taking the cross-sectional and longitudinal results together there is some evidence that higher volumes and intensity of LTPA may be associated with reduced risk of hip fracture later in life, while the highest levels may be associated with increased odds of a fracture, not isolated to hip, during mid-life.

While previous prospective studies of LTPA and hip fracture risk have also reported that higher levels of activity are associated with lower fracture risk,^22–24^ ours is the first study to model LTAV continuously using restricted cubic splines, allowing the shape of the dose-response relationship, including the attenuation of the inverse association at higher activity volumes, to be examined. UKB has previously been described as a ‘healthy cohort’,^25^ consequently it contains more participants reporting high volumes of activity than previous studies.^22^ This was also reflected in the relatively high absolute activity volumes represented by the upper exposure quintiles, allowing associations at these higher volumes to be examined separately.

When activity was examined according to intensity, the strongest inverse associations with hip fracture risk were observed for vigorous activity. However, UKB does not ask about how activity is accumulated, making it hard to examine causal pathways between intensity, impact and in particular BMD in this cohort. In terms of possible mechanisms by which increased physical activity, in particular vigorous activity, might reduce hip fracture risk, there is good evidence from studies using BMD as an outcome, that higher volumes and intensity of weight bearing activity can lead to beneficial changes in the density and architecture of femoral bone, with a resultant increase in strength.^5^ However, it is unclear whether generally healthy adults can naturally reach these osteogenic loading patterns and achieve the resultant beneficial effects while undertaking habitual LTPA, particularly in later life. Accelerometer studies suggest that very few community-dwelling older women experience vertical impacts of sufficient magnitude to stimulate bone adaptation during daily living.^26^ While hip BMD was measured several years after the initial physical activity questionnaire in approximately 20% of UKB participants, there were insufficient numbers to explore its role in mediating the relationship we observed between physical activity and hip fracture. That said, adjustment for baseline estimated heel BMD as a proxy measure for hip BMD did not materially change the relationship of physical activity with hip fracture risk.

An alternative mechanistic explanation is that physical activity in mid-life is representative of the broader health status of the individual, with physical fitness embodying other characteristics such as muscle strength, agility, balance and co-morbidity that a) impact on the ability to undertake higher levels of physical activity, and b) are closely associated with sarcopenia, and in turn falls and fracture risk in later life. In particular grip strength and walking pace have previously been found to be strong indicators of physical capability and are independent predictors of mortality beyond common sociodemographic markers, as well as being key components of commonly used sarcopenia definitions.^20,21^ In our analysis, we found that adjustment for grip strength and falls had little effect on the association with future hip fracture. However, when we included self-reported walking pace in our models, the association for all but vigorous activity was largely attenuated. Walking pace is considered to be a marker of overall functional capability and health,^27^ and may capture shared elements of fitness, physical reserve and physical function with attained activity volume. As our measures of activity and walking pace were measured concurrently it is not possible to determine in our data whether walking pace is acting as a confounder or mediator of the relationship with fracture. Furthermore, the inverse association of vigorous activity with hip fracture risk only partly attenuated after adjustment for walking pace, implying that some aspects of vigorous activity act independently of walking pace.

The findings of this study are broadly consistent with advice from the National Osteoporosis Guideline Group (NOGG), and the Royal Osteoporosis Societies position statement *“Strong, Steady and Straight”* that intensity/type of activity as well as volume plays an important role in fracture prevention.^28,29^ NOGG also draw a clear distinction between evidence for changes to bone density and fracture outcomes concluding that the evidence that exercise reduces fracture risk remains unclear. This study helps address this gap, strengthening the evidence for a role of activity in fracture prevention and adding to the body of evidence for more detailed quantification of amount, intensity and type of activity required for benefit.

### Strengths and Limitations

A key strength of this study is the large sample size which allowed stratification by sex, as well as detailed analysis of different intensities of activity, as well as volume. A limitation of our work is that although population based, UKB represents a selected sample which may limit generalisability of findings given the relative health and homogeneous ethnicity of the cohort. Furthermore, causal inferences cannot be made from physical activity measured at a single time point in mid-life, as we do not know how activity behaviours changed between baseline assessment and fracture, or whether mid-life activity is an indicator of subsequent activity. Physical activity was measured subsequently by accelerometery in a subset of UKB participants, however, the shorter follow-up time from accelerometer assessment limits the number of hip fracture events available for analyses of this rare outcome, and the accelerometer-derived measures are not directly equivalent to the self-reported LTPA measured examined here. Residual confounding by factors not measured, or incompletely measured, in UKB also cannot be excluded. The use of self-reported measures raised the risk of measurement and reporting errors.

## Conclusions

Mid-life LTPA was associated with lower long-term hip fracture risk, with the lowest risk observed at approximately one hour of total LTPA per day. Approximately 60–90 minutes of vigorous activity per week was associated with the lowest hazard of fracture, independent of walking and moderate activity, with partial attenuation at higher volumes. These findings suggest that both the volume and intensity of mid-life physical activity may be important in relation to subsequent hip fracture risk with the inverse associations attenuating at the highest activity volumes.

## Data Availability

The data analysed in this study are available from the UK Biobank website with approved access. Full code and outputs related to this analysis is available at GitHub (https://github.com/RollsCf/UKB_PA_Fracture.git)

## Disclosures and Declarations

### Funding

This work was supported by the Wellcome Trust through a GW4-CAT-HP fellowship to C.R. (grant no. 316291/Z/24/Z). The views in this article are those of the authors and do not necessarily reflect the views of the Wellcome Trust. BGF is supported by a Wellcome Trust Early Career Award (316390/Z/24/Z). HD is funded by NIHR Exeter BRC and HRC

### Ethical approval

This study did not require ethical approval. Ethical approval for the UKB cohort was provided by NHS Northwest Multicentre Research Ethics Committee (21/NW/0157).

### Conflicts of Interest

Catherine Rolls, Marcus Munafo, Helen Dawes, Jon Tobias and Emma Clark declare no conflicts of interest in relation to this piece of work. BGF has received honoraria from UCB and Calico Life Sciences and done consulting for Relation Therapeutics.

## Acknowledgments

The authors would like to thank the participants of UK Biobank for their contribution to this ongoing cohort study and the Wellcome Trust for supporting this research

## Supplementary Methods

Methods S1. Changes to the analyses plan

Supplementary Methods S1
|  |  |
| --- | --- |
| Plan | We will use two different outcome variables: hip fracture and wrist fracture. While there are limitations of using the HES dataset, as it only captures data from those admitted to hospital, preparatory work done by our group suggests there is good validity of HES data for hip and wrist fracture. |
| Change | We restricted our prospective analysis to hip fracture. This was a pragmatic decision due to the number of analyses being run and time limitations. We elected to focus on the fracture outcome with the largest clinical significance |
| Impact | Moderate |
| Plan | Exposure variable: Planned analysis<br>Continuous MET-minutes per week for:<br>moderate activity<br>vigorous activity<br>walking<br>moderate-to-vigorous physical activity (MVPA)<br>total physical activity (sum of moderate, vigorous, and walking MET-minutes) |
| Change | We did not evaluate MVPA in these analyses as we did not feel it added anything over and above the other intensity measures. |
| Impact | Low |
| Plan | Additional exploratory sensitivity analyses adjusting for heel BMD, grip strength, walking speed and falls history were not in the pre-registered plan. |
| Change | Added after completion of the original statistical analysis plan to explore potential mechanistic explanations and assess the robustness of the observed associations. These analyses were specified before interpretation of the study results. |
| Impact | Moderate |

## Supplementary Tables

**Table S1:**
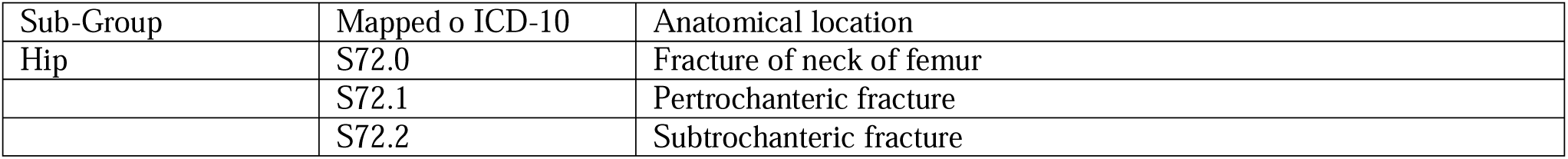
International Classification of Disease Version 10 (ICD10) codes relating to hip fracture types used as the outcome.

| Sub-Group | Mapped o ICD-10 | Anatomical location |
| --- | --- | --- |
| Hip | S72.0 | Fracture of neck of femur |
|  | S72.1 | Pertrochanteric fracture |
|  | S72.2 | Subtrochanteric fracture |

**Table S2.** Response rate and degree of missing data.

| Variable | Answered | Answered (%) | Missing | Missing (%) |
| --- | --- | --- | --- | --- |
| Ethnicity | 425737 | 99.45 | 2338 | 0.55 |
| Days/week moderate activity | 406360 | 94.93 | 21715 | 5.07 |
| Days/week walking | 419567 | 98.01 | 8508 | 1.99 |
| Days/week vigorous activity | 406654 | 95.00 | 21421 | 5.00 |
| Moderate activity volume (MET-min/week) | 368265 | 86.03 | 59810 | 13.97 |
| Vigorous activity volume (MET-min/week) | 383931 | 89.69 | 44144 | 10.31 |
| Leisure walking volume (MET-min/week) | 372667 | 87.06 | 55408 | 12.94 |
| Leisure time physical activity volume (MET-min/week) | 332403 | 77.65 | 95672 | 22.35 |
| Education | 415537 | 97.07 | 12538 | 2.93 |
| Townsend deprivation index | 427501 | 99.87 | 574 | 0.13 |
| Weight (kg) | 425715 | 99.45 | 2360 | 0.55 |
| Height (cm) | 425909 | 99.49 | 2166 | 0.51 |
| Body mass index (kg/m<sup>2</sup>) | 425441 | 99.38 | 2634 | 0.62 |

MET = Metabolic Equivalent Task Unit, Kg = Kilograms, cm = centimetres, m = meters, min= minutes

**Table S3.** Missing data comparison.

| Variable | Overall | Activity data available | Activity data missing | SMD |
| --- | --- | --- | --- | --- |
| Sex, n (%) | NA | NA | NA | 0.099 |
| Female | 234,155 (55%) | 175,130 (53%) | 59,025 (63%) |  |
| Male | 190,572 (45%) | 155,692 (47%) | 34,880 (37%) |  |
| Age (years), mean (SD) | 54.7 (7.2) | 54.4 (7.3) | 55.5 (7.1) | 0.145 |
| Ethnicity, n (%) | NA | NA | NA | 0.025 |
| Asian | 8,723 (2.1%) | 5,902 (1.8%) | 2,821 (3.0%) |  |
| Black or other | 12,876 (3.0%) | 9,114 (2.8%) | 3,762 (4.0%) |  |
| Mixed | 2,714 (0.6%) | 2,095 (0.6%) | 619 (0.7%) |  |
| White | 398,985 (94%) | 312,807 (95%) | 86,178 (92%) |  |
| Missing | 1,429 | 904 | 525 |  |
| Townsend deprivation index, mean (SD) | -1.3 (3.1) | -1.4 (3.0) | -0.8 (3.3) | 0.171 |
| Missing | 569 | 446 | 123 |  |
| Education level, n (%) | NA | NA | NA | 0.013 |
| School education or less | 348,211 (84%) | 273,382 (84%) | 74,829 (85%) |  |
| College or vocational training | 39,189 (9.5%) | 30,982 (9.5%) | 8,207 (9.4%) |  |
| University degree or above | 25,737 (6.2%) | 21,032 (6.5%) | 4,705 (5.4%) |  |
| Missing | 11,590 | 5,426 | 6,164 |  |
| Weight (kg), mean (SD) | 78.2 (16.2) | 78.3 (16.0) | 77.8 (16.6) | 0.029 |
| Missing | 1,960 | 1,170 | 790 |  |
| Height (cm), mean (SD) | 168.7 (9.3) | 169.2 (9.3) | 166.7 (9.1) | 0.279 |
| Missing | 1,771 | 1,044 | 727 |  |

<sup>2</sup>SMD = standardised mean difference. Values <0.1 indicate negligible differences, 0.1–0.2 small differences, and >0.2 meaningful imbalance. n= number % = percentage SD = standard deviation, cm = centimetres

**Table S4:** Pooled Cox regression models for the association between leisure-time physical activity volume and hip fracture, including tests for non-linearity and sex interaction.

| Outcome | N | Events | Analysis | Result | Estimate | P_value |
| --- | --- | --- | --- | --- | --- | --- |
| Hip fracture | 324284 | 2674 | Continuous model | Unadjusted | HR 0.963 (95% CI 0.938, 0.988) | 0.004 |
| Hip fracture | 324284 | 2674 | Continuous model | Minimally adjusted | HR 0.938 (95% CI 0.914, 0.964) | <0.001 |
| Hip fracture | 324284 | 2674 | Continuous model | Fully adjusted | HR 0.938 (95% CI 0.914, 0.964) | <0.001 |
| Hip fracture | 324284 | 2674 | Non-linearity | Linear vs restricted cubic spline LRT | Chi-square 13.43, df 2 | 0.001 |
| Hip fracture | 324284 | 2674 | Interaction | Sex × LTPA volume LRT | Chi-square 1.39, df 3 | 0.707 |

**Note:** Hazard ratios (HRs) are presented with 95% confidence intervals from Cox proportional hazards models. The likelihood ratio test (LRT) compared the fit of the fully adjusted linear model with a restricted cubic spline model to assess evidence of non-linearity, and compared models with and without a sex × physical activity interaction term to assess effect modification by sex. A significant LRT P value indicates that the more complex model provided a significantly better fit to the data.

**Table S5:** Proportional hazards tests for pooled fully adjusted spline models.

| Outcome | Model | Variable | Chi_square | df | P_value |
| --- | --- | --- | --- | --- | --- |
| Hip fracture | Fully adjusted spline | Physical activity | 3.851 | 3 | 0.278 |
| Hip fracture | Fully adjusted spline | Sex | 18.121 | 1 | <0.001 |
| Hip fracture | Fully adjusted spline | Ethnicity | 3.917 | 3 | 0.271 |
| Hip fracture | Fully adjusted spline | Height | 12.518 | 1 | <0.001 |
| Hip fracture | Fully adjusted spline | Weight | 0.209 | 1 | 0.648 |
| Hip fracture | Fully adjusted spline | Townsend deprivation index | 0.952 | 1 | 0.329 |
| Hip fracture | Fully adjusted spline | Education | 0.726 | 2 | 0.695 |
| Hip fracture | Fully adjusted spline | Global | 40.843 | 12 | <0.001 |

**Note:** Proportional hazards assumptions were assessed using Schoenfeld residuals (cox.zph). A significant P value indicates evidence against the proportional hazards assumption for the corresponding covariate or for the model overall (Global). The pooled models demonstrated evidence of non-proportional hazards, with the strongest evidence observed for sex, which motivated subsequent sex-stratified analyses.

**Table S6.**
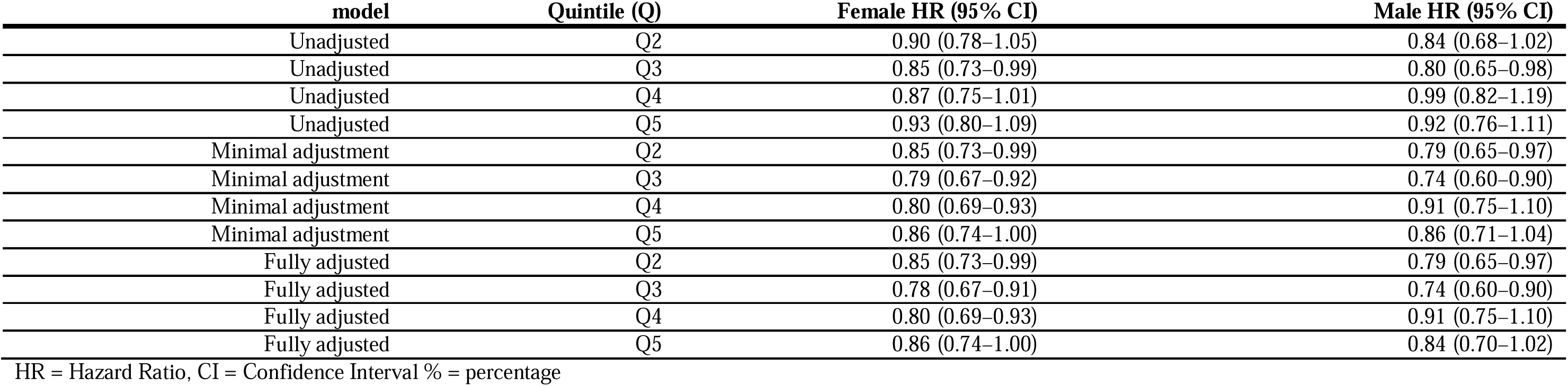
Hazard ratios for incident hip fracture across sex-specific quintiles of LTPAv.

**Table S7:**
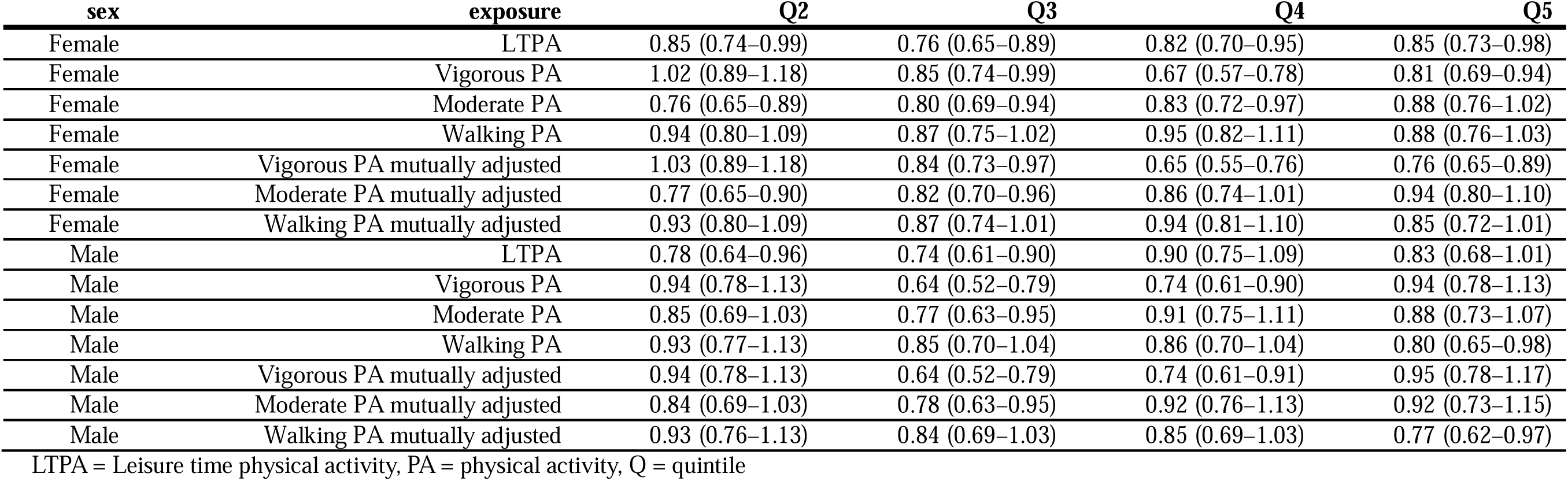
Hazard ratios for hip fracture according to physical activity intensity quintiles.

**Table S8:** Sensitivity analyses of PA and incident hip fracture with additional adjustment for falls, grip strength, walking pace and heel bone mineral density.

| Exposure | Marker | Analysis | Female Q2 | Female Q3 | Female Q4 | Female Q5 | Male Q2 | Male Q3 | Male Q4 | Male Q5 |
| --- | --- | --- | --- | --- | --- | --- | --- | --- | --- | --- |
| Total PA | Falls | Fully adjusted | 0.85 (0.73–0.99) | 0.78 (0.67–0.91) | 0.80 (0.69–0.93) | 0.86 (0.74–1.00) | 0.79 (0.65–0.97) | 0.74 (0.60–0.90) | 0.91 (0.76–1.11) | 0.85 (0.70–1.02) |
| Total PA | Falls | Additionally adjusted | 0.85 (0.73–0.99) | 0.78 (0.67–0.91) | 0.80 (0.69–0.93) | 0.85 (0.73–0.99) | 0.81 (0.66–0.99) | 0.75 (0.61–0.92) | 0.93 (0.77–1.12) | 0.84 (0.70–1.02) |
| Total PA | Grip strength | Fully adjusted | 0.85 (0.73–0.99) | 0.78 (0.67–0.91) | 0.80 (0.68–0.93) | 0.86 (0.74–1.00) | 0.80 (0.65–0.98) | 0.74 (0.61–0.91) | 0.92 (0.76–1.11) | 0.85 (0.70–1.02) |
| Total PA | Grip strength | Additionally adjusted | 0.87 (0.75–1.01) | 0.80 (0.69–0.94) | 0.82 (0.70–0.95) | 0.89 (0.76–1.03) | 0.82 (0.67–1.00) | 0.77 (0.62–0.94) | 0.96 (0.79–1.17) | 0.90 (0.74–1.09) |
| Total PA | Walking pace | Fully adjusted | 0.85 (0.73–0.99) | 0.78 (0.67–0.91) | 0.80 (0.69–0.93) | 0.86 (0.74–1.00) | 0.79 (0.65–0.97) | 0.74 (0.60–0.90) | 0.91 (0.75–1.10) | 0.84 (0.70–1.02) |
| Total PA | Walking pace | Additionally adjusted | 0.97 (0.84–1.13) | 0.92 (0.79–1.08) | 0.96 (0.82–1.12) | 1.05 (0.90–1.23) | 0.89 (0.73–1.10) | 0.86 (0.70–1.05) | 1.07 (0.88–1.30) | 1.01 (0.83–1.22) |
| Total PA | Heel BMD | Fully adjusted | 0.90 (0.74–1.10) | 0.81 (0.66–0.99) | 0.84 (0.68–1.02) | 0.93 (0.76–1.13) | 0.74 (0.57–0.97) | 0.71 (0.54–0.92) | 0.97 (0.76–1.23) | 0.86 (0.67–1.09) |
| Total PA | Heel BMD | Additionally adjusted | 0.91 (0.75–1.11) | 0.84 (0.68–1.03) | 0.87 (0.71–1.06) | 0.96 (0.79–1.17) | 0.78 (0.60–1.01) | 0.76 (0.58–0.99) | 1.06 (0.84–1.35) | 0.93 (0.73–1.19) |
| Walking PA | Falls | Fully adjusted | 0.94 (0.80–1.09) | 0.87 (0.75–1.02) | 0.95 (0.82–1.11) | 0.88 (0.76–1.03) | 0.94 (0.77–1.14) | 0.85 (0.70–1.04) | 0.86 (0.71–1.05) | 0.81 (0.66–0.99) |
| Walking PA | Falls | Additionally adjusted | 0.94 (0.81–1.09) | 0.87 (0.75–1.02) | 0.95 (0.82–1.10) | 0.87 (0.75–1.02) | 0.95 (0.78–1.16) | 0.86 (0.71–1.05) | 0.87 (0.71–1.06) | 0.80 (0.65–0.98) |
| Walking PA | Grip strength | Fully adjusted | 0.94 (0.81–1.09) | 0.87 (0.75–1.02) | 0.95 (0.82–1.11) | 0.88 (0.76–1.03) | 0.94 (0.77–1.14) | 0.86 (0.70–1.05) | 0.86 (0.71–1.05) | 0.80 (0.66–0.99) |
| Walking PA | Grip strength | Additionally adjusted | 0.95 (0.81–1.10) | 0.89 (0.76–1.03) | 0.97 (0.83–1.13) | 0.90 (0.77–1.05) | 0.95 (0.78–1.15) | 0.86 (0.71–1.05) | 0.87 (0.72–1.06) | 0.83 (0.67–1.01) |
| Walking PA | Walking pace | Fully adjusted | 0.94 (0.80–1.09) | 0.87 (0.75–1.02) | 0.95 (0.82–1.11) | 0.88 (0.76–1.03) | 0.93 (0.77–1.13) | 0.85 (0.70–1.04) | 0.86 (0.70–1.04) | 0.80 (0.65–0.98) |
| Walking PA | Walking pace | Additionally adjusted | 1.06 (0.91–1.24) | 1.02 (0.87–1.19) | 1.14 (0.97–1.32) | 1.05 (0.90–1.23) | 1.02 (0.84–1.24) | 0.96 (0.79–1.18) | 0.98 (0.80–1.20) | 0.93 (0.75–1.14) |
| Walking PA | Heel BMD | Fully adjusted | 1.02 (0.84–1.25) | 0.91 (0.74–1.12) | 1.01 (0.82–1.23) | 1.00 (0.81–1.22) | 0.97 (0.75–1.25) | 0.90 (0.69–1.16) | 0.94 (0.73–1.21) | 0.82 (0.63–1.06) |
| Walking PA | Heel BMD | Additionally adjusted | 1.02 (0.84–1.25) | 0.92 (0.75–1.13) | 1.02 (0.83–1.25) | 1.02 (0.83–1.25) | 0.99 (0.77–1.28) | 0.93 (0.72–1.20) | 0.97 (0.75–1.25) | 0.85 (0.65–1.10) |
| Moderate PA | Falls | Fully adjusted | 0.76 (0.65–0.89) | 0.80 (0.69–0.94) | 0.83 (0.72–0.97) | 0.88 (0.76–1.02) | 0.85 (0.69–1.04) | 0.77 (0.63–0.95) | 0.92 (0.76–1.11) | 0.89 (0.73–1.08) |
| Moderate PA | Falls | Additionally adjusted | 0.77 (0.65–0.90) | 0.81 (0.69–0.94) | 0.84 (0.72–0.97) | 0.87 (0.76–1.01) | 0.86 (0.71–1.06) | 0.79 (0.64–0.97) | 0.93 (0.77–1.13) | 0.88 (0.73–1.07) |
| Moderate PA | Grip strength | Fully adjusted | 0.76 (0.65–0.89) | 0.80 (0.69–0.94) | 0.83 (0.72–0.97) | 0.87 (0.75–1.01) | 0.84 (0.69–1.03) | 0.78 (0.64–0.95) | 0.92 (0.76–1.11) | 0.88 (0.73–1.07) |
| Moderate PA | Grip strength | Additionally adjusted | 0.77 (0.66–0.91) | 0.82 (0.71–0.96) | 0.86 (0.74–1.00) | 0.90 (0.78–1.04) | 0.88 (0.71–1.07) | 0.81 (0.66–1.00) | 0.97 (0.80–1.17) | 0.95 (0.78–1.15) |
| Moderate PA | Walking pace | Fully adjusted | 0.76 (0.65–0.89) | 0.80 (0.69–0.94) | 0.83 (0.72–0.97) | 0.88 (0.76–1.02) | 0.85 (0.69–1.03) | 0.77 (0.63–0.95) | 0.91 (0.75–1.11) | 0.88 (0.73–1.07) |
| Moderate PA | Walking pace | Additionally adjusted | 0.86 (0.73–1.01) | 0.93 (0.79–1.08) | 0.98 (0.84–1.14) | 1.04 (0.89–1.20) | 0.96 (0.78–1.18) | 0.90 (0.73–1.10) | 1.07 (0.88–1.30) | 1.03 (0.85–1.25) |
| Moderate PA | Heel BMD | Fully adjusted | 0.75 (0.61–0.92) | 0.71 (0.58–0.88) | 0.89 (0.73–1.07) | 0.85 (0.70–1.02) | 0.92 (0.71–1.19) | 0.78 (0.60–1.02) | 0.96 (0.75–1.23) | 0.92 (0.72–1.19) |
| Moderate PA | Heel BMD | Additionally adjusted | 0.76 (0.62–0.94) | 0.74 (0.60–0.91) | 0.91 (0.75–1.11) | 0.86 (0.71–1.04) | 0.95 (0.73–1.23) | 0.82 (0.63–1.07) | 1.03 (0.81–1.33) | 0.97 (0.76–1.25) |
| Vigorous PA | Falls | Fully adjusted | 1.03 (0.89–1.18) | 0.85 (0.74–0.99) | 0.67 (0.57–0.78) | 0.81 (0.69–0.94) | 0.94 (0.79–1.13) | 0.64 (0.52–0.79) | 0.74 (0.60–0.90) | 0.94 (0.78–1.13) |
| Vigorous PA | Falls | Additionally adjusted | 1.03 (0.89–1.18) | 0.85 (0.74–0.99) | 0.67 (0.57–0.79) | 0.81 (0.70–0.94) | 0.95 (0.79–1.14) | 0.65 (0.53–0.80) | 0.75 (0.61–0.91) | 0.94 (0.78–1.13) |
| Vigorous PA | Grip strength | Fully adjusted | 1.02 (0.89–1.18) | 0.86 (0.74–0.99) | 0.67 (0.57–0.78) | 0.81 (0.69–0.94) | 0.95 (0.79–1.14) | 0.64 (0.52–0.79) | 0.75 (0.61–0.91) | 0.94 (0.78–1.14) |
| Vigorous PA | Grip strength | Additionally adjusted | 1.02 (0.89–1.18) | 0.88 (0.76–1.02) | 0.69 (0.59–0.81) | 0.84 (0.72–0.98) | 0.96 (0.80–1.16) | 0.67 (0.55–0.82) | 0.79 (0.65–0.96) | 1.00 (0.83–1.21) |
| Vigorous PA | Walking pace | Fully adjusted | 1.02 (0.89–1.18) | 0.85 (0.74–0.99) | 0.67 (0.57–0.78) | 0.81 (0.69–0.94) | 0.94 (0.78–1.13) | 0.64 (0.52–0.79) | 0.74 (0.61–0.90) | 0.94 (0.78–1.13) |
| Vigorous PA | Walking pace | Additionally adjusted | 1.03 (0.90–1.18) | 0.96 (0.82–1.11) | 0.76 (0.65–0.89) | 0.94 (0.81–1.10) | 0.97 (0.81–1.17) | 0.74 (0.60–0.91) | 0.86 (0.70–1.05) | 1.09 (0.90–1.32) |
| Vigorous PA | Heel BMD | Fully adjusted | 1.16 (0.96–1.39) | 0.87 (0.71–1.06) | 0.68 (0.55–0.84) | 0.89 (0.73–1.08) | 0.86 (0.68–1.09) | 0.56 (0.43–0.73) | 0.72 (0.56–0.92) | 0.91 (0.71–1.15) |
| Vigorous PA | Heel BMD | Additionally adjusted | 1.16 (0.96–1.39) | 0.90 (0.74–1.10) | 0.71 (0.57–0.87) | 0.95 (0.78–1.16) | 0.87 (0.69–1.10) | 0.60 (0.46–0.79) | 0.80 (0.62–1.03) | 1.03 (0.81–1.30) |

SR = Self-reported, BMD = Bone Mineral Density, Quintiles show Hazard Ratio and 95% confidence interval

## Supplementary Figures

**Figure S1:**
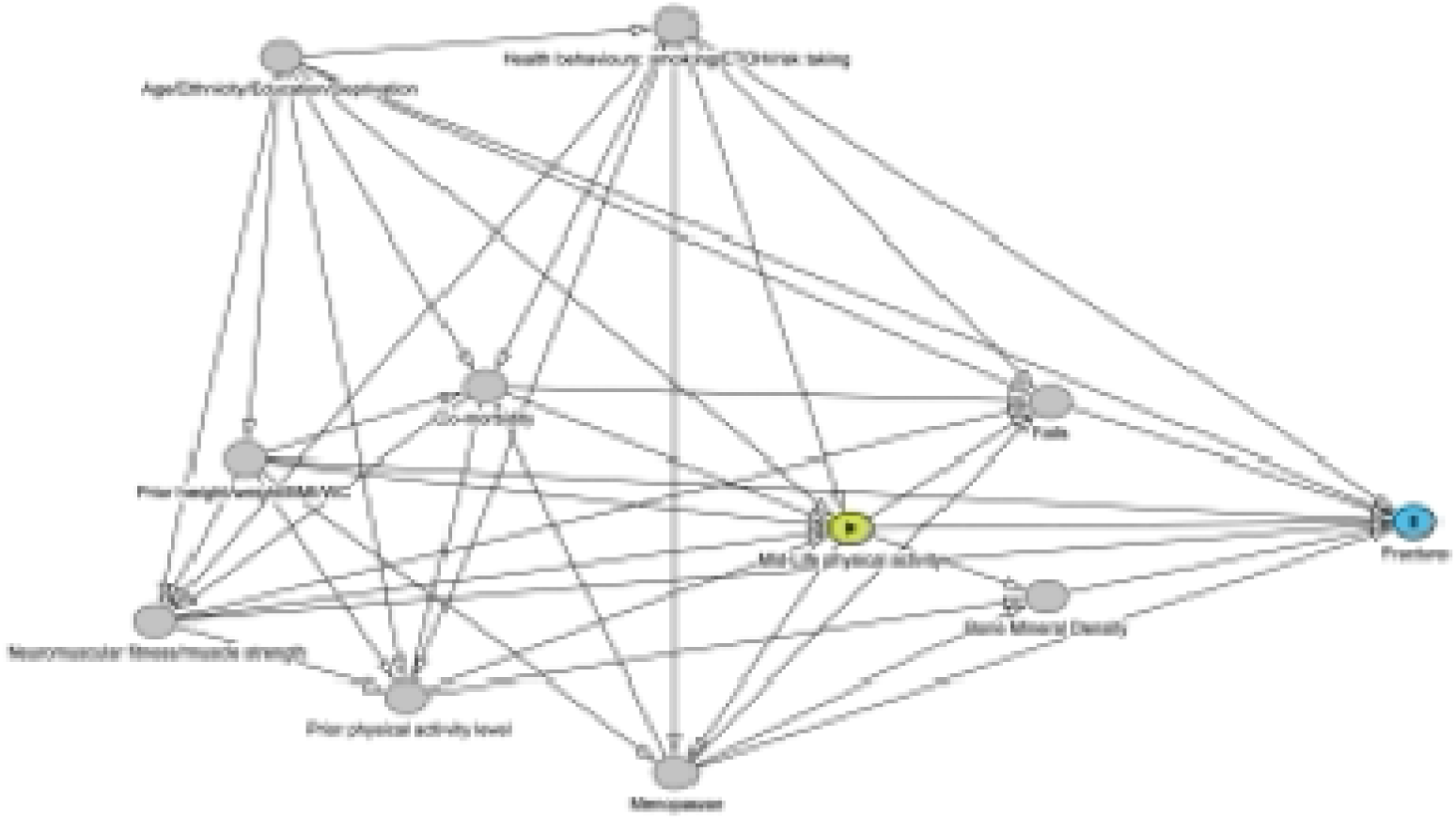
Directed Acyclic Graph of the possible pathways from mid-life physical activity to fracture. The directed acyclic graph (DAG) summarises the hypothesised relationships between mid-life physical activity (exposure), fracture (outcome), and measured covariates. Arrows represent assumed causal relationships based on prior evidence. Age, ethnicity, education/deprivation, health behaviours, prior physical activity, body size, neuromuscular fitness, comorbidity, and menopause were considered potential confounders. Falls and bone mineral density were considered potential mediators of the association between physical activity and fracture and were not included in the primary adjustment set. The green circle with a triangle denotes the exposure, and the blue circle the outcome.

**Figure S2.**
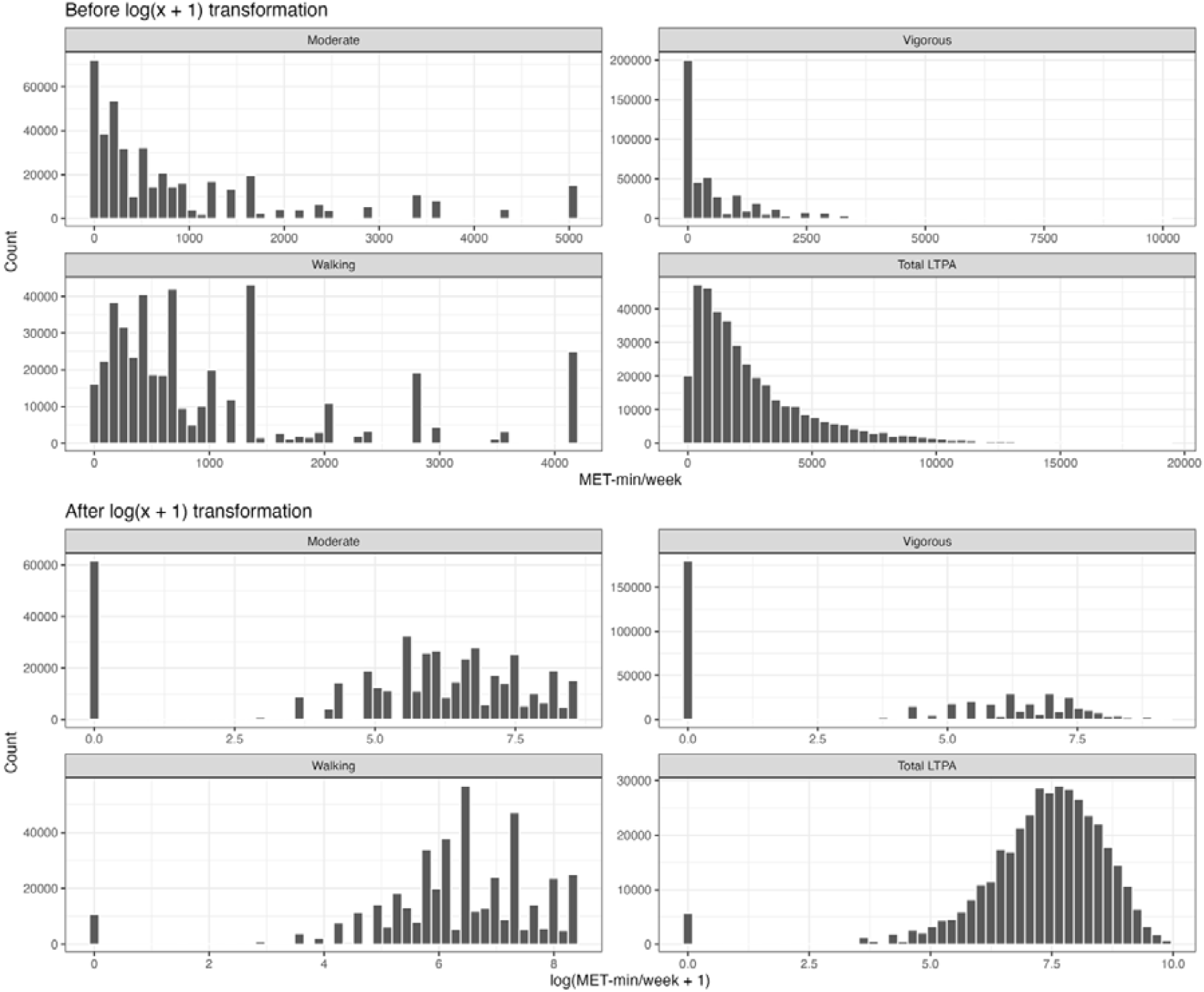
Distribution of physical activity variables before and after log(MET-min/week + 1) transformation. Histograms show the distributions of moderate-intensity activity, vigorous-intensity activity, walking, and LTPA_V_ (MET-min/week) before (top panels) and after (bottom panels) log(MET-min/week + 1) transformation. The transformation reduced the strong positive skew observed in the original distributions and improved their suitability for modelling as continuous exposures in restricted cubic spline analyses.

**Figure S3:**
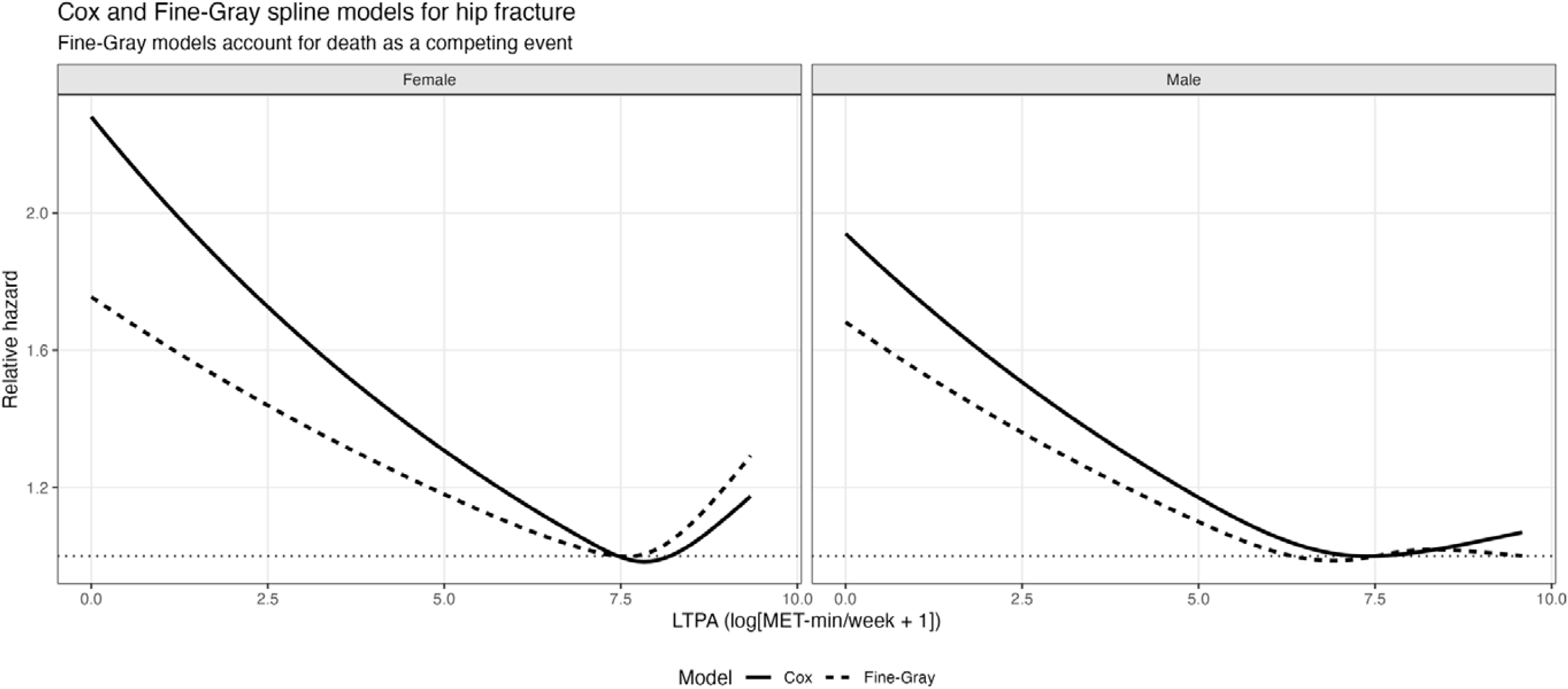
**Cox and Fine-Gray cubic spline models for the association between Leisure Time Physical Activity (LTPA) volume and hip fracture, stratified by sex**. Solid lines represent the cox model and dashed lines represent the estimates from Fine-Gray models. Leisure-time physical activity (LTPA) volume was modelled continuously as log-transformed MET-min/week using restricted cubic splines. Models were fully adjusted for ethnicity, height, weight, Townsend deprivation index and education level.

**Figure S4.**
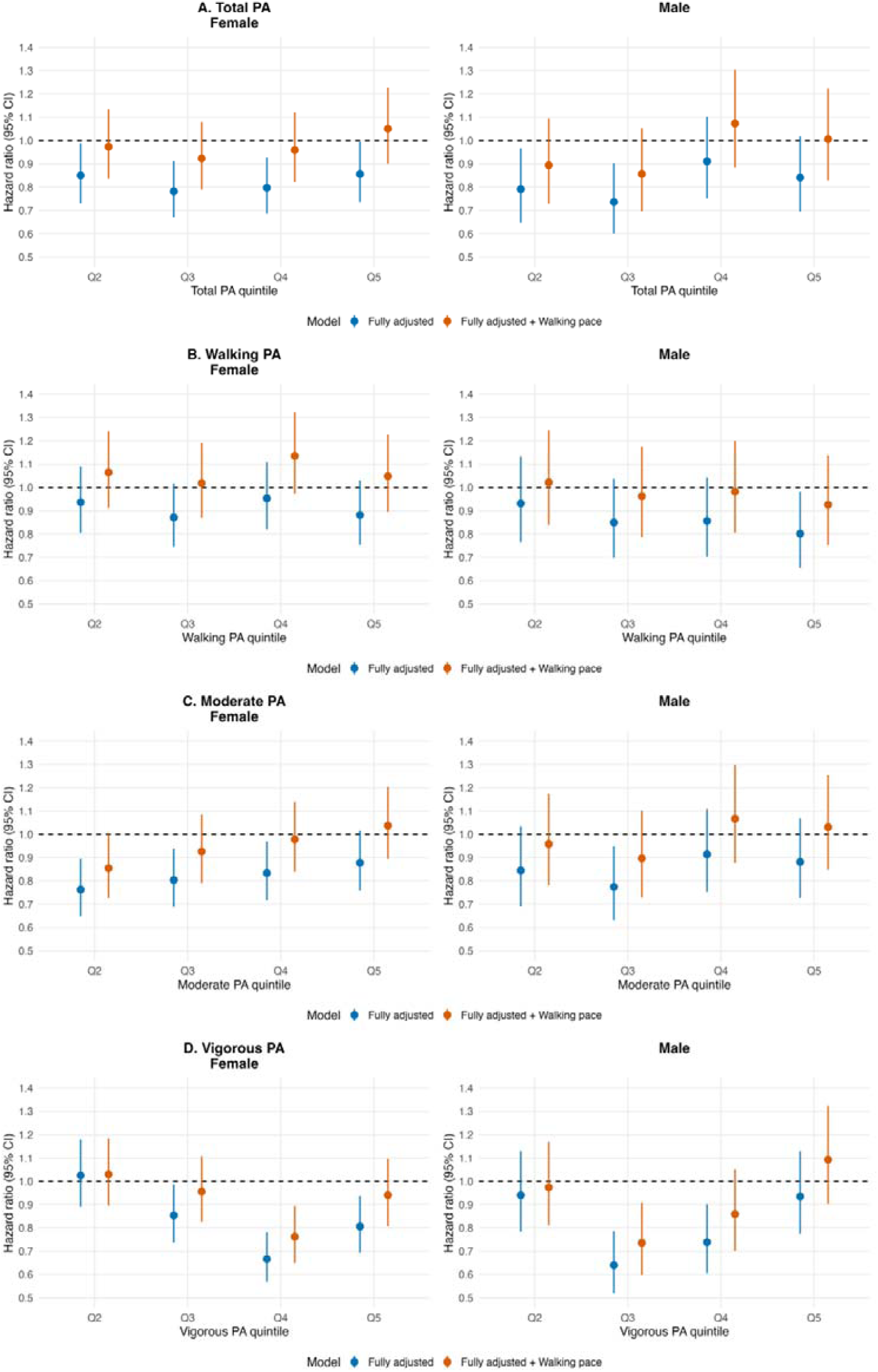
Hazard ratios (HRs) and 95% confidence intervals for hip fracture according to quintiles of walking, moderate-intensity, and vigorous-intensity physical activity, stratified by sex. Estimates are from fully adjusted Cox proportional hazards compared to additional adjustment for self-reported walking pace. Quintile 1 (lowest activity level) was used as the reference category.

